# Integrating multi-modal omics to identify therapeutic atherosclerosis pathways for coronary heart disease

**DOI:** 10.1101/2024.12.11.24318833

**Authors:** Sophie C. de Ruiter, Marion van Vugt, Chris Finan, Diederick E. Grobbee, Dominique P.V. de Kleijn, Gerard Pasterkamp, Hester M. den Ruijter, Ernest Diez Benavente, Sanne A.E. Peters, A. Floriaan Schmidt

## Abstract

**Introduction:** Urinary metabolism breakdown products reflect metabolic changes in atherosclerosis-relevant tissues and may contain relevant therapeutic leads. We integrated data on urinary metabolism breakdown products, plasma proteins, atherosclerotic plaque tissue, and single-cell expression to identify druggable metabolic pathways for coronary heart disease (CHD).

**Methods:** Mendelian randomisation was employed to interrogate findings from independent genome-wide association studies on 954 urinary metabolism breakdown products, 1,562 unique proteins, and 181,522 CHD cases, establishing directionally concordant associations. Using the Athero-Express Biobank, concordant plasma proteins were linked to plaque vulnerability using protein and mRNA expression in plaque. Single-cell RNA sequencing data obtained from carotid plaque samples were used to test for differential expression of concordant proteins across plaque cell types.

**Results:** In total, 29 urinary metabolism breakdown products associated with CHD, predominantly originating from amino acid metabolism (n=12) or unclassified origin (n=9). We identified 113 plasma proteins with directionally concordant associations with these urinary metabolism breakdown products and CHD. Of the 110 proteins available in plaque, 16 were associated with plaque vulnerability. This included positive control proteins targeted by drugs indicated for CHD, such as CAH1 (targeted by aspirin), IL6R (targeted by tocilizumab), and AT1B2 (targeted by digoxin), as well as two potential repurposing opportunities C1S (targeted by C1-esterase inhibitor and sutimlimab) and CATH (targeted by bortezomib).

**Conclusion:** We have identified amino acid metabolism as an important contributing pathway to CHD risk and prioritised 16 proteins relevant for CHD with involvement in atherosclerotic plaques, providing important insights for drug development.

## Introduction

Atherosclerosis is characterised by the development of lesions accumulating lipoproteins and inflammatory cells in the arterial wall. As atherosclerosis progresses, plaque accumulates and may progress into high-risk vulnerable plaques. Such plaques are characterized by a thin fibrous cap, a large lipid core and high levels of inflammatory cells(1, 2), making them particularly prone to rupture. Plaque rupture can lead to rapid thrombus formation blocking the blood flow to the heart muscle, potentially leading to acute clinical events such as coronary heart disease (CHD), which remains a leading cause of burden and death globally.

Metabolic disorders, such as metabolic syndrome (the co-occurrence of hypertension, hypercholesterolemia, diabetes, and obesity), are associated with a metabolic, pro-inflammatory, and pro-thrombotic state which are key drivers of atherosclerosis(3), reflecting the close interrelationship between immune response and metabolic homeostasis, where dysfunction in one system can adversely affect the other(4). Plasma proteins are markers of disease and targets of most drug compounds. Previous research has already linked plaque phenotypes to circulating proteins, identifying reduced plasminogen activator inhibitor levels in patients with an intermediate coronary artery disease risk profile, for example(5).

Urinary metabolism breakdown products, which are the end-products of metabolism excreted in urine, reflect metabolic changes occurring in atherosclerosis-relevant tissues (e.g., body fat, liver, the arterial wall). These urinary breakdown products may provide key insights in the metabolic alterations associated with the progression of atherosclerosis to CHD. While urinary breakdown products are potentially important markers of disease, they are not directly actionable as they have been excreted from the body. On the contrary, plasma proteins are actionable targets for therapeutic intervention and therefore the integration of data on urinary metabolites and plasma proteins identifies relevant metabolic pathways and potential drug targets, offering a comprehensive approach to understanding and intervening in the progression of CHD.

In the current study, we used Mendelian randomisation (MR) to identify novel metabolic pathways for CHD by leveraging genetic data on urinary metabolism breakdown products (954 urinary metabolism breakdown products), plasma proteins (1,562 unique plasma proteins), and CHD (181,522 cases). The fixed nature of genotypes, established during gametogenesis(6, 7) allows genetic variants to be used as instrumental variables in MR, which is therefore robust against the presence of confounding bias and potential reverse causation. In the context of cardiovascular disease, this approach has been empirically validated(8–11). Plaque involvement was established by using data from carotid endarterectomy patients from the Athero-Express (AE) Biobank on protein expression, mRNA expression, and single-cell RNA sequencing. Finally, the potential druggability of these prioritised proteins was evaluated through linkage to ChEMBL.

## Methods

### Genetic data

Genetic associations with 954 urinary metabolism breakdown product values were available from a genome-wide association study (GWAS) conducted on 1,627 individuals using a non-targeted mass spectrometry Metabolon assay(12). Metabolism breakdown products were grouped based on their metabolic origins, which led to nine metabolism classes: amino acid metabolism, carbohydrate metabolism, cofactor and vitamin metabolism, energy metabolism, lipid metabolism, nucleotide metabolism, peptide metabolism, xenobiotic metabolism, or unclassified metabolism. Metabolism breakdown products with an origin in xenobiotic metabolism are breakdown products from substances not naturally produced by the human body, such as drugs, pollutants, and synthetic compounds. Unclassified metabolism breakdown products represent potentially novel biomarkers, as they have not yet been related to an established metabolic origin. These breakdown products were noted for their recurring chromatographic and spectral properties and have been assigned unique COMP-IDs and/or CHEM-IDs (**Appendix Table S1**), allowing for identification and comparison in future studies and by other analysis platforms. We used eight GWAS on genetic associations with plasma protein values, detailed in the **Supplemental Methods**, and a GWAS on genetic associations with incident or prevalent CHD (181,522 cases among 1,165,690 participants from European ancestry) obtained from Aragam *et al.*(13).

### Mendelian randomization analyses

Three MR analyses were conducted: 1) genome-wide MR to identify urinary metabolism breakdown products associated with CHD through upstream effects in plasma (step 1 in **Figure 1, Appendix Table S1**), 2) *cis*-MR to identify plasma proteins affecting values of urinary metabolism breakdown products (step 2 in **Figure 1**), and 3) *cis*-MR to identify plasma proteins associated with CHD (step 3 in **Figure 1**). For the genome-wide MR, genetic variants were selected from across the genome, while the *cis*-MR utilized genetic variants selected from a 200 kilobase pair window around and within the protein encoding gene. In all MRs, variants were selected based on an exposure F-statistic of at least 24 and a minimal minor allele frequency of 0.01. The selected variants were clumped to a linkage disequilibrium (LD) r-squared of 0.30, using a random sample of 5,000 unrelated UK Biobank participants(14). The *cis*-MR analyses (steps 2-3 in **Figure 1**) were conducted per individual protein GWAS, where the largest sample size GWAS was used if protein measurements overlapped in different studies.

**Figure 1.**
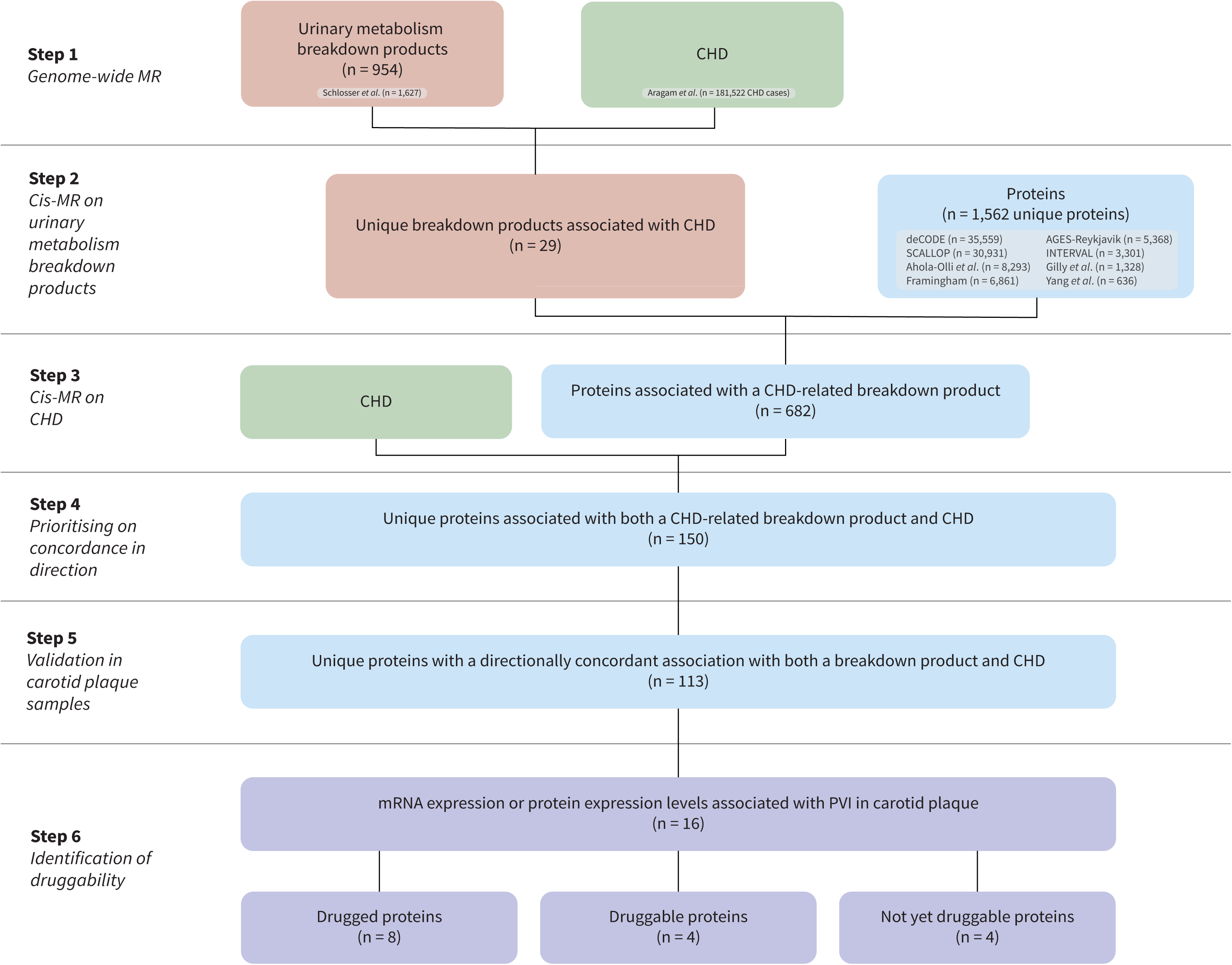
Flowchart of the main steps of this study. N.B. The first three steps are conducted using Mendelian randomisation, based on indicated GWAS data for CHD, urinary metabolism breakdown products, and plasma proteins. Directionally concordance is explained in Figure 2 and the Methods section. Measurements on mRNA and protein expression are available from carotid plaque samples from the Athero-Express Biobank. Druggability of plasma proteins is determined using ChEMBL and the British National Formulary. Abbreviations: CHD = coronary heart disease, MR = Mendelian randomisation, PVI = plaque vulnerability index.

MR analyses were conducted using generalized least squares (GLS) implementations of the inverse variance weighted (IVW) estimator, as well as MR-Egger estimator, known for its robustness to pleiotropy(15). The GLS implementation was used to additionally correct for residual LD, which optimized estimator precision(16). To reduce the potential for horizontal pleiotropy, variants with large leverage statistics (larger than three times the mean leverage) and/or outlier statistics (Chi-square larger than 10.83) were excluded. We discarded analyses with fewer than six variants, ensuring we had sufficient data to accurately model the exposure effects. To further minimize the potential of horizontal pleiotropy, we applied a model selection framework identifying the MR model (IVW or MR-Egger) that is most supported by the available data(17).

### Triangulation of evidence through consideration of concordant CHD effects

We integrated findings from the three distinct MR analyses to triangulate evidence on the effects of urinary metabolism breakdown products and proteins on CHD. This involved 1) identifying urinary metabolism breakdown products affecting CHD, 2) identifying proteins associated with these urinary metabolism breakdown products, and 3) focussing on the subset of proteins with an effect on CHD that was directionally concordant with the effect of a urinary metabolism breakdown product on CHD and the protein on the same breakdown product. For example, if a higher value of a urinary metabolism breakdown product reduced CHD risk, and a protein increased the value of this urinary breakdown product, the protein effect was considered directionally concordant if higher protein values reduced CHD risk. We describe concordance in direction as a triangulated association (**Figure 2**). Given that potential horizontal pleiotropy acts through distinct pathways in each analysis (e.g., pre-translational pleiotropy in *cis*-MR versus pre- or post-translational horizontal pleiotropy in genome-wide MR(18)), focusing on results with directional concordance ensures identification of a robust subset of results with likely limited residual bias due to horizontal pleiotropy.

**Figure 2.**
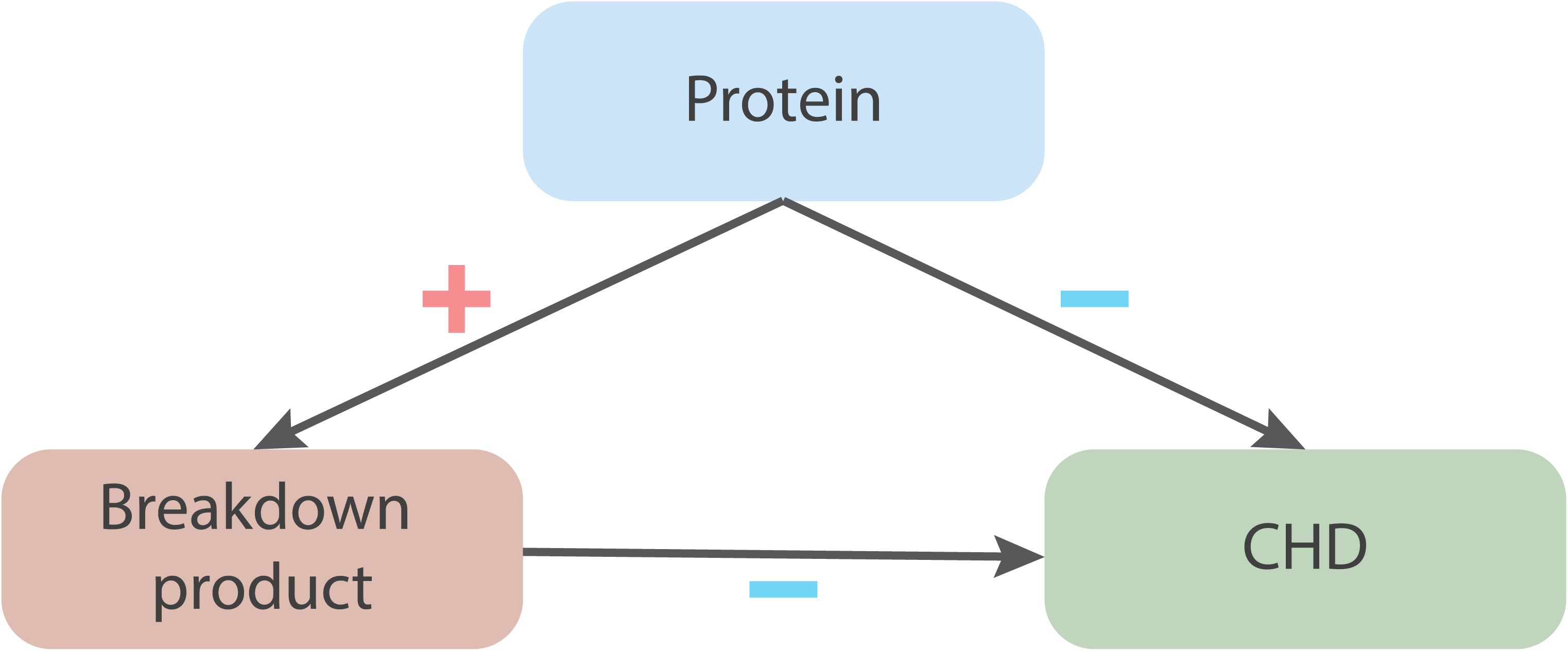
Example of a triangulated association with directional concordance between a protein and its effects on urinary metabolism breakdown product values and CHD. N.B. The plus symbol (in red) represents a positive association. The minus symbol (in blue) represents a negative association (a risk-decreasing effect). Directional concordance is achieved when the direction of effects aligns consistently, whether increasing or decreasing.

### Athero-Express Biobank atherosclerotic plaques

Next, the Athero-Express (AE) Biobank data was used to prioritise findings on atherosclerotic plaque involvement. Patients enrolled in the AE Biobank were at risk of CHD, as well as other atherosclerosis associated diseases such as ischemic stroke, with carotid plaques from these patients providing a relevant marker of generalised atherosclerosis (54–58). Specifically, the AE contains samples of carotid endarterectomy patients, where we identified potential associations between protein (194 patients) or mRNA (632 patients) expression in plaque and the plaque vulnerability index. In addition, single-cell RNA sequencing data (4948 cells and 46 patients) were analysed to study cellular plaque expression. Please see **Supplemental Methods** for the relevant details, and **Appendix Table S2** for AE patients characteristics. The performed study is in line with the Declaration of Helsinki, and informed consent was provided by all study participants after approval for this study by the medical ethical committees of the various hospitals (University Medical Center, Utrecht, The Netherlands, and St. Antonius Hospital, Nieuwegein, The Netherlands) was obtained.

### Associations with plaque vulnerability

We tested for associations between both protein expression and mRNA expression levels of MR-prioritised proteins and plaque vulnerability index using a linear model, with protein expression available for 1,500 proteins and mRNA expression available from 55,105 transcripts, all measured in atherosclerotic plaques.

### Determining cellular expression using single-cell RNA sequencing

Single-cell RNA sequencing was used to explore whether genes coding for MR-prioritised proteins where differentially expressed across plaque cell types, providing important information on the mechanisms driving atherosclerosis. A Wilcoxon rank-sum test was used to compare the expression in a single cell type to expression in the remaining cell types. The following cell types were considered: dendritic cells, endothelial cells I (ECs I), endothelial cells II (ECs II), foam cells, inflammatory macrophages, mast cells, memory B-cells, monocytes, natural killer cells (NK-cells), plasma B-cells, resident macrophages, smooth muscle cells (SMCs), and T-cells. Additionally, differential expression across broader clusters of cell types (structural cells, innate immune cells, and adaptive immune cells) was assessed using the Wilcoxon rank-sum test (**Supplemental Methods**).

### Annotations, effect estimates and multiple testing

Proteins are referred to using their Uniprot label(19) and genes are referred to using the Ensembl label in italicised font. MR results are presented as mean differences (MD) for urinary metabolism breakdown products, or odds ratios (OR) for CHD, representing the effect of one standard deviation (SD) increase of the exposure (i.e., either urinary metabolism breakdown products or plasma protein value). Associations of the plaque vulnerability index with protein expression levels or mRNA expression levels are presented as MD in normalized count per unit increase in the plaque vulnerability index. All effect estimates are accompanied by 95% confidence intervals (CIs) and p-values. Metabolism breakdown product effects on CHD were filtered for a multiplicity corrected p-value threshold of 1.37×10LJLJ based on the 365 principal components that were needed to explain 90% of the variance in the urinary metabolism breakdown products (**Appendix Figure S1**). Protein MR effect estimates of the metabolism breakdown product association were evaluated against a multiplicity corrected p-value threshold of 1.10×10^-6^ based on the number of tested proteins (n=1,562) and metabolism breakdown products (n=29). The multiplicity corrected p-value threshold was 7.33×10^-6^ for the MRs of protein on CHD based on the number of tested proteins (n=682). For analyses of associations of protein and mRNA expression levels with plaque vulnerability, p-values were adjusted using the Benjamini-Hochberg method(20) with a false discovery rate threshold of 0.1. For differential expression testing using single-cell RNA sequencing data, we used a nominal p-value threshold of 0.05.

### Druggability of prioritised proteins

We identified plasma proteins targeted by approved drugs (drugged proteins), as well as plasma proteins targeted by a developmental drug (druggable proteins) based on ChEMBL. For the drugged and druggable proteins, compound indications and side-effects were extracted from ChEMBL and the British National Formulary (BNF).

### Partial replication of protein associations with cardiac traits

Due to the availability of eight proteomics GWAS, some studies measured the same proteins, which we used to replicate the associations with CHD risk. Replication was sought using a nominally significant p-value of 0.05 or smaller and considering the effect direction of the triangulated analysis. In addition, a more stringent p-value threshold of 6.94×10^-4^ (0.05 divided by the number of proteins that were available in more than one study) was used for more conservative replication.

## Results

### Urinary metabolism breakdown products associating with CHD

Using Mendelian randomisation, we evaluated 954 urinary metabolism breakdown products to investigate their association with CHD (**Appendix Table S1**). Out of these, 29 were associated with CHD (**Appendix Table S3,** step 1 in **Figure 1, Figure 3**). Twelve of these metabolism breakdown products originated from amino acid metabolism. For example, higher values of N-acetyltyrosine increased the risk of CHD (OR 1.03 per one standard deviation increase, 95%CI 1.02; 1.04), while higher values of 3-methylglutaconate decreased the risk of CHD (OR 0.95 per one standard deviation increase, 95%CI 0.93; 0.97). Other associated urinary metabolism breakdown products originate from various metabolic processes: one from energy metabolism, four from lipid metabolism, and three from xenobiotic metabolism. Finally, nine urinary metabolism breakdown products of unclassified metabolic origin were associated with CHD, representing potential novel metabolic pathways.

**Figure 3.**
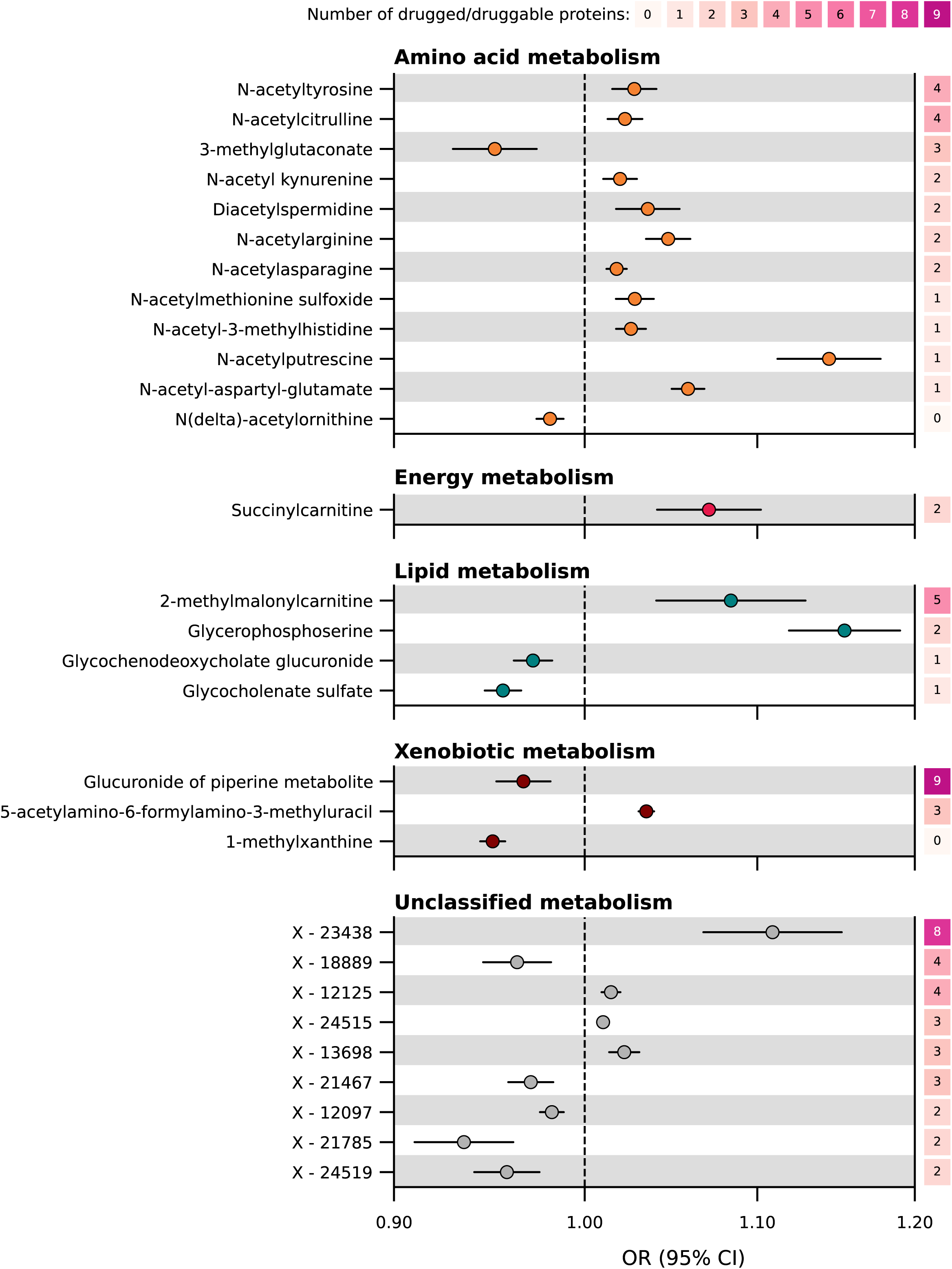
Associations of urinary metabolism breakdown products with CHD, presented per metabolism class. N.B. Point estimates represent associations between urinary metabolism breakdown products and coronary heart disease (CHD), obtained from genome-wide Mendelian randomisation. The metabolism classes (amino acid, energy, lipid, xenobiotics, and unclassified) represent the metabolic origins of the breakdown product. The right y-axis indicates the number of drugged and druggable proteins associated with the metabolism breakdown product and CHD, where druggability is sourced from the British National Formulary and ChEMBL. Genetic associations with 954 urinary metabolism breakdown products were obtained from Schlosser *et al.* (n=1,627)(12). Genetic associations with CHD were obtained from Aragam *et al*. (181,522 CHD cases)(13). For more detailed information, please refer to the Methods section and **Appendix Tables S1-S2**. Abbreviations: CI = confidence interval, OR = odds ratio.

### Triangulated proteins concordant with urinary metabolism breakdown products and CHD

We used Mendelian randomisation to determine associations between plasma proteins and the 29 urinary metabolism breakdown products identified as being associated with CHD. We identified 682 proteins associating with at least one of these 29 urinary metabolism breakdown products (**Appendix Figure S2)**. Among these, 113 proteins were identified with a directionally concordant association with CHD (steps 2-4 in **Figure 1, Appendix Figure S3, Appendix Table S4**). Please see, **Supplemental Results** and **Appendix Tables S5-S6** for MR analyses replicating 83% of these findings in smaller independent GWAS.

### MR-prioritised proteins and plaque vulnerability index

Of the 113 MR-prioritised proteins, 36 were available and detectable in the AE protein expression data measured in plaque samples (194 patients, **Appendix Table S2** for characteristics) and 10 proteins were associated with increased plaque vulnerability: A2M, AKR7A2, APOF, C1S, CA1, COMT, CTSD, CTSH, NAGK, and PLTP (**Appendix Figure S4, Appendix Table S7**). Data on mRNA expression in plaques was available for 110 genes coding for MR-prioritised proteins (632 patients, **Appendix Table S2** for characteristics), identifying an additional 6 proteins where mRNA expression was associated with plaque vulnerability: FER, IL6RA, ATF6B, AT1B2, SWP70, and SIG14 (**Figure 4, Appendix Figure S5, Appendix Table S8**).

**Figure 4.**
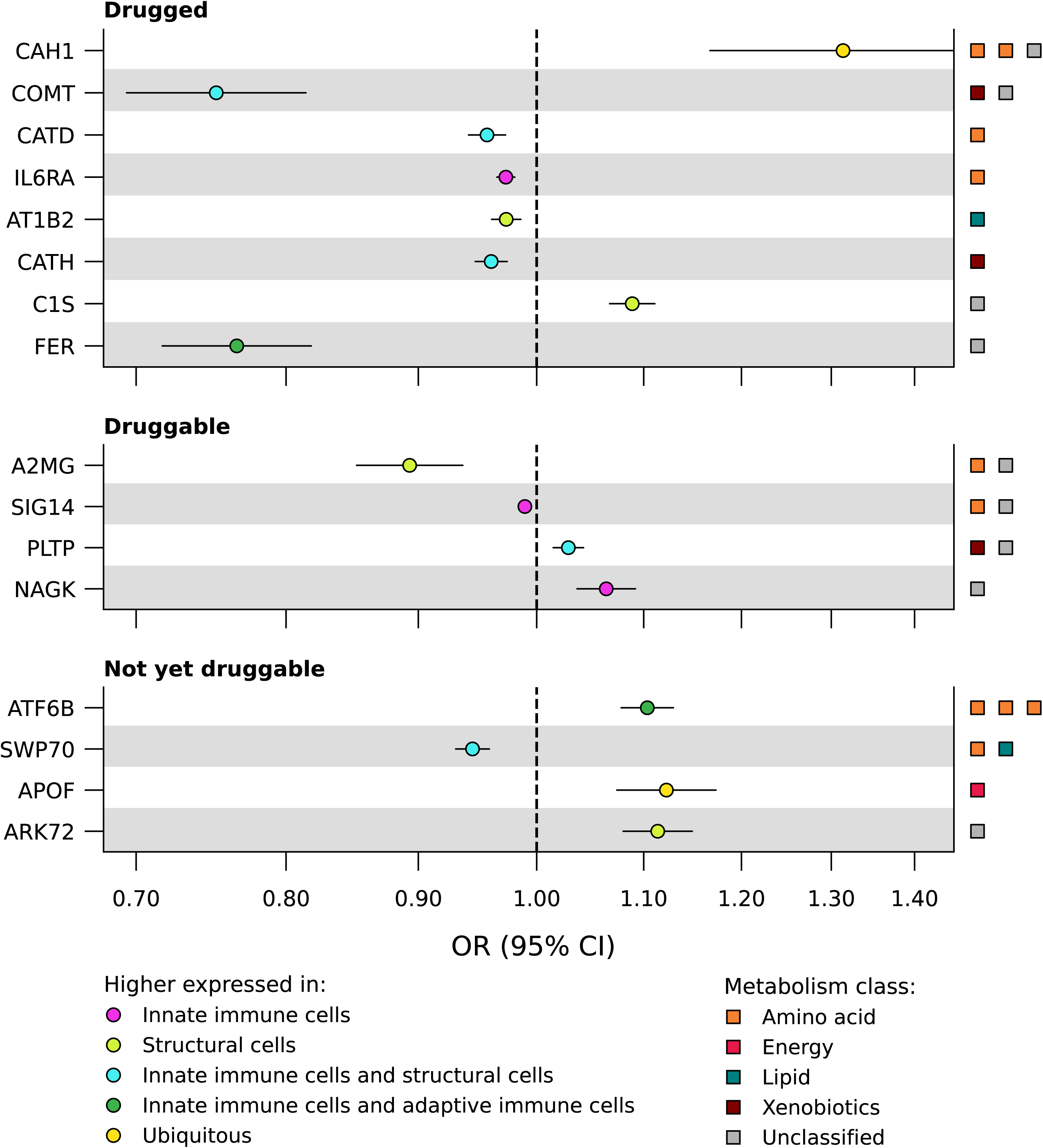
Associations of prioritised proteins with CHD and plaque vulnerability in atherosclerotic plaque. N.B. Point estimates represent associations between plasma proteins and coronary heart disease (CHD), obtained from *cis* Mendelian randomisation. Druggability is based on ChEMBL and the British National Formulary, distinguishing between proteins targeted by approved drug (drugged proteins, top panel), proteins targeted by a drug in development (druggable proteins, middle panel) and not yet druggable proteins (bottom panel). Differential expression of genes coding for prioritised proteins is determined across cell types based on single-cell RNA sequencing data obtained from Athero-Express patients (4948 cells, 46 patients). The metabolism classes of the metabolism breakdown products affected by the protein are indicated on the right y-axis. Genetic associations with CHD were obtained from Aragam *et al*. (181,522 CHD cases)(13). For a more detailed description, please refer to the Methods section and the **Supplemental Methods**. Full numerical results can be found in **Appendix Table S4**. Abbreviations: CI = confidence interval, OR = odds ratio.

### Single-cell RNA sequencing of MR-prioritised proteins

To understand the cellular origin of the proteins, we additionally explored which genes encoding prioritised proteins were differentially expressed across cells in carotid plaques. For this, we used single-cell RNA sequencing data on over 4,900 cells available for 105 of our MR-prioritised proteins including all 16 proteins associated with plaque vulnerability. We identified 87 genes that showed to be enriched in one or more plaque cell types, which included 14 genes encoding for the proteins associated with plaque vulnerability (**Appendix Table S9, Figure 5**). For example, the gene *ATP1B2*, coding the protein AT1B2, was higher expressed in smooth muscle cells as compared to other plaque cell types (**Figure 5, Appendix Figure S6, Appendix Table S9**). The gene *ATF6B*, coding for the protein ATF6B, was higher expressed in T-cells as compared to other plaque cell types. Next, we used broader clusters of cell types, namely structural cells, innate immune cells, and adaptive immune cells. We found three (IL6RA, SIG14, and NAGK) out of 16 plaque vulnerability-associated proteins to be higher expressed in innate immune cells as compared to the other two clusters of cell types, while four (AT1B2, C1S, A2MG, and ARK72) were higher expressed in structural cells, five (COMT, CATD, CATH, PLTP, and SWP70) were higher expressed in both innate immune and structural cells and two (FER and ATF6B) were higher expressed in both innate immune cells and adaptive immune cells (**Figure 4**). Two proteins (CAH1 and APOF) were ubiquitous (i.e., no higher expression in any of the three clusters).

**Figure 5.**
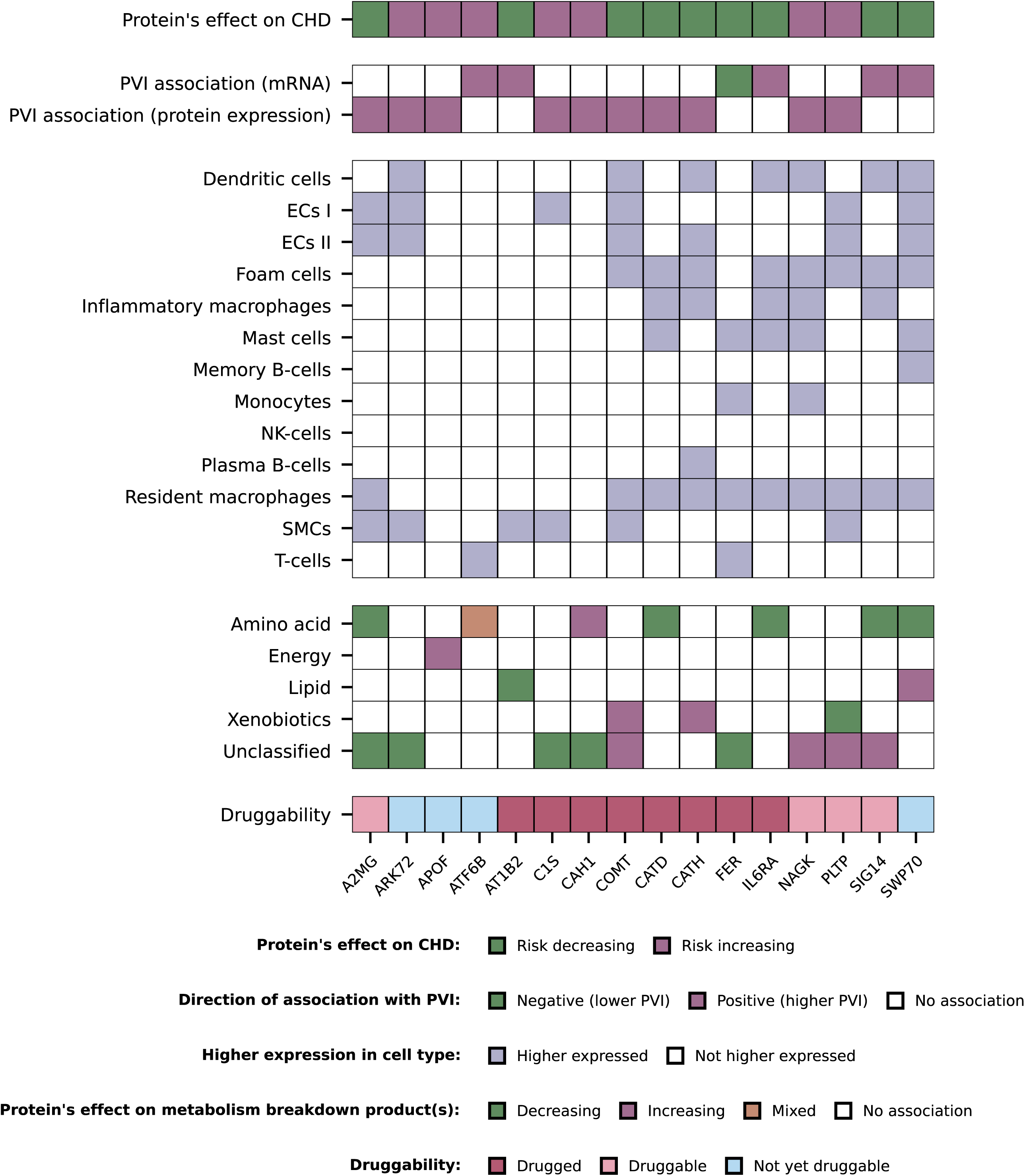
Matrix of prioritised proteins associated with CHD and plaque vulnerability in atherosclerotic plaque. N.B. Each column represents a prioritised protein associated with coronary heart disease (CHD) and plaque vulnerability. The gene names corresponding to each protein are presented in **Table 1**. The rows present the following: row 1, direction of the protein’s association with CHD as obtained by Mendelian randomisation; row 2-3, association of the gene’s mRNA or protein expression in plaque with plaque vulnerability, obtained from the Athero-Express Biobank; row 4-16, higher expression in a plaque cell type as compared to expression in all other plaque cell types, obtained from single-cell RNA sequencing in the Athero-Express Biobank; row 17-21, direction of the protein’s association with metabolism breakdown product(s) presented per metabolism class, based on Mendelian randomisation; row 22, druggability status, distinguishing proteins targeted by approved drug (drugged proteins) from proteins targeted by a drug in development (druggable proteins) and not yet druggable proteins, based on ChEMBL and the British National Formulary. Full numerical results can be found in **Appendix Tables S4**, **S7-S9**. Abbreviations: CHD = coronary heart disease, ECs = endothelial cells, NK-Cells = natural killer cells, PVI = plaque vulnerability index, SMCs = smooth muscle cells.

**Table 1.**
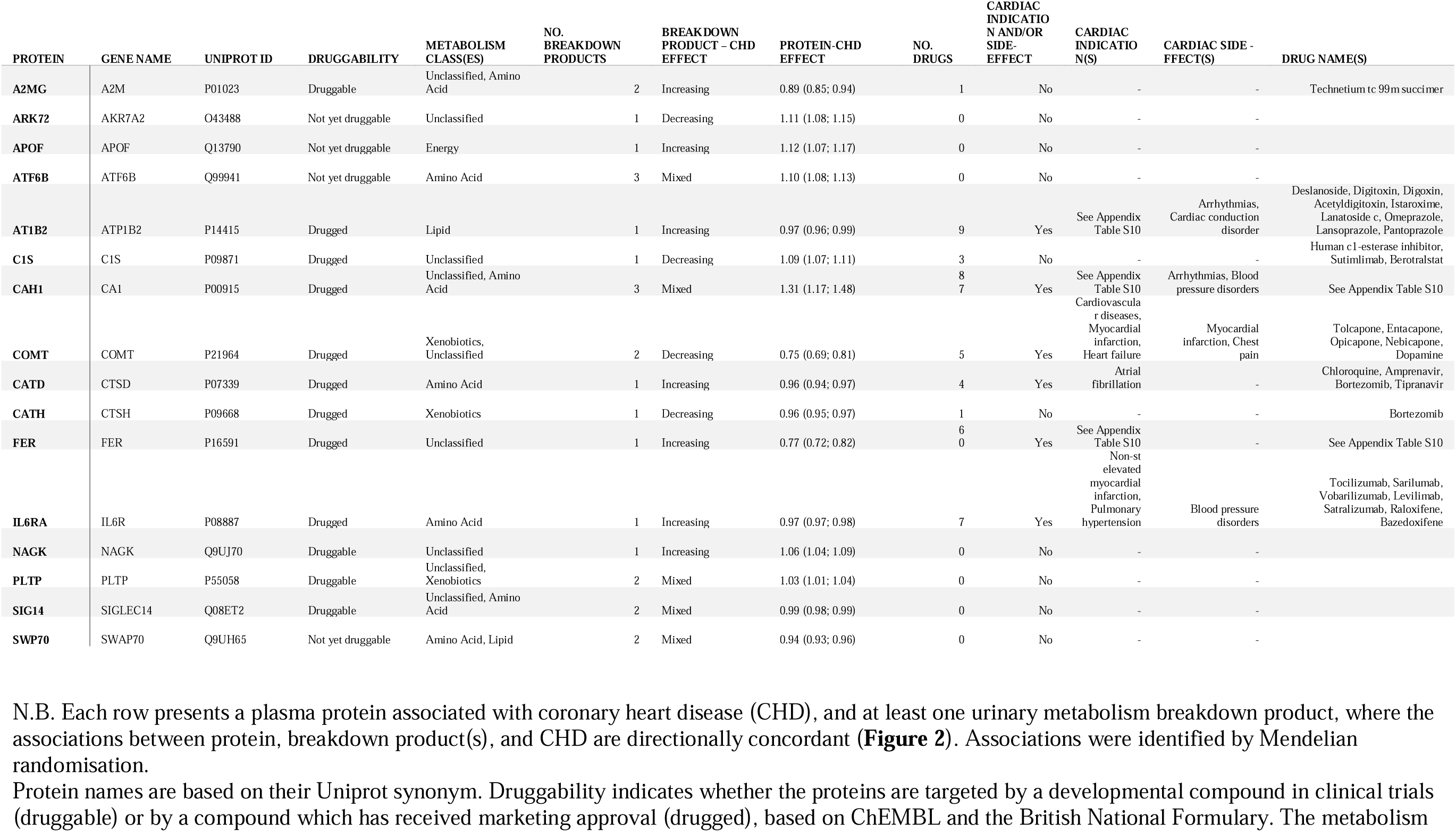
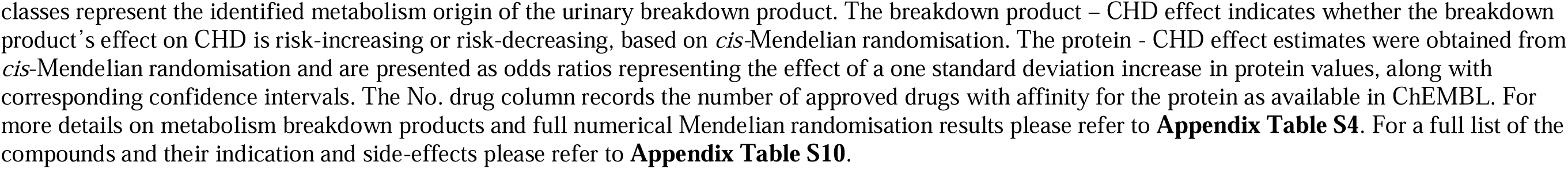
Positive controls and repurposing opportunities: prioritised proteins related to plaque vulnerability.

### Druggability of proteins

The set of 16 proteins associated with plaque vulnerability included eight drugged proteins, four druggable proteins, and four not yet druggable proteins (step 6 in **Figure 1, Figure 4-5)**. These proteins predominantly associated with amino acid breakdown products and unclassified breakdown products. Six drugged proteins (AT1B2, CAH1, CATD, COMT, FER, and IL6RA) were targeted by a drug with a cardiac indication and/or side-effect (**Table 1**, **Figure 4, Appendix Tables S10-S11**). The protein AT1B2 (targeted as part of a protein complex group through digoxin which is contraindicated in HCM) decreased the risk of CHD (OR 0.97, 95%CI 0.96; 0.99). Higher values of CAH1 (targeted by methocarbamol which has registered side-effects including blood pressure disorders, and by dopamine and aspirin indicated for myocardial infarction and coronary artery disease) increased the risk of CHD (OR 1.31, 95%CI 1.17; 1.48). Higher values of COMT (targeted by the drugs tolcapone and entacapone belonging to the class of COMT-inhibitors indicated for Parkinson’s disease which have side-effects including chest pain and increased risk of CHD) decreased the risk of CHD (OR 0.75, 95%CI 0.69; 0.81). Higher values of IL6RA, interleukin-6 receptor subunit alpha, decreased the risk of CHD (OR 0.97, 95%CI 0.97; 0.98). IL6RA is targeted by the interleukin-6 receptor antagonist tocilizumab indicated for auto-immune diseases, which is also in phase 2 development for CHD(21) and has registered side-effects of blood pressure disorders. Higher values of CATD (targeted by chloroquine indicated for malaria and cancer treatment, and in development for atrial fibrillation) decreased the risk of CHD (OR 0.96, 95%CI 0.94; 0.97). Higher values of FER (targeted by everolimus indicated in cancer and in development for acute coronary syndrome) decreased the risk of CHD (OR 0.77, 95%CI 0.72; 0.82).

KLKB1 and PGFRB are two proteins directionally concordantly associated with unclassified and amino acid metabolism breakdown products and CHD. Both are targeted by drugs with cardiac indications or side-effects, but neither is associated with plaque vulnerability. Higher values of KLKB1, targeted by the inhibitor lanadelumab (in phase 2 development for hypotension during hemodialysis), decreased the risk of CHD (OR 0.97, 95%CI 0.95; 0.98). Similarly, higher values of PGFRB, targeted by the inhibiting cancer drugs regorafenib and dasatinib (which have known side-effects including CHD, QT interval prolongation and blood pressure disorders), decreased the risk of CHD (OR 0.98, 95%CI 0.98; 0.99).

## Discussion

In this study, we used Mendelian randomisation to assess relationships between 954 urinary metabolism breakdown products, 1,562 proteins and CHD, and we linked protein and mRNA expression levels in carotid plaque to plaque vulnerability. This was done to identify potential biomarkers of altered metabolism informative of CHD risk and plasma proteins which may be used as targets for intervention in these metabolic pathways. Through our multi-layered approach we were able to prioritise 16 proteins associated with plaque vulnerability and CHD risk, where 83% of these CHD associations were independently replicated. The majority of these proteins were associated with urinary metabolism breakdown products from amino acid metabolism or unclassified origins.

Altered amino acid metabolism in cardiovascular disease has been observed before(38–41). For example, the kynurenine pathway has been linked to the pathogenesis of atherosclerosis by modulating inflammation, oxidative stress, and endothelial function(42). Its breakdown products were identified as biomarkers of major cardiovascular events(43, 44), with previous studies showing the therapeutic potential of targeting this pathway to target vascular inflammation and plaque formation(45). We were able to link N-acetylkynurenine to the druggable proteins SIG14 and A2MG. SIG14 is member of the sialic acid-binding immunoglobulin-type lectins (Siglecs) family, which is involved in immune regulation and has been studied as drug target for various diseases including asthma, cancer, and autoimmune diseases(46). A2MG is a broad-spectrum protease inhibitor with an anti-inflammatory role and it has a dual role in coagulation(47, 48), which we now link to plaque vulnerability.

The 16 prioritised proteins included two proteins with a known indication or side-effect on CHD: CAH1 and COMT. CAH1, is targeted by aspirin which is prescribed to reduce the risk of blood clots(22) in people at a high risk of (subsequent) cardiovascular events. COMT is affected by various drugs, including the Parkinson’s disease drugs entacapone and tolcapone which is known to increase the risk of myocardial infarction through effects on blood pressure, arteriosclerosis, and aortic stenosis(23).

These positive control findings confirmed our analysis was able to identify known drug targets effect for CHD. Additionally, we identified four drugged proteins (AT1B2, CATD, FER, and IL6RA) which are in development for CHD or CHD-related diseases. IL6R inhibiting drugs (e.g., tocilizumab) are currently being repurposed for treatment of CHD(8, 11), where the phase 3 CANTOS trial evaluating canakinumab, inhibiting IL-1 which is upstream of IL6 and IL6R, showed clinical effectiveness in preventing CHD(24). This provides additional support for our approach as we rediscovered known drug target effects for CHD.

The drugged proteins C1S and CATH were identified as potential repurposing candidates for treatment of CHD and atherosclerosis. CATH is targeted by Bortezomib which is indicated for multiple myeloma, and due to side-effects such as thrombocytopenia, and neutropenia, not directly relevant to consider for CHD management. C1S on the other hand is targeted by inhibiting drugs (e.g., berotralstat and human c1-esterase inhibitor) indicated for conditions such as angioedema. C1S activates the classical pathway of the complement system, contributing to inflammation and endothelial dysfunction, which promotes atherosclerosis(25, 26). The here identified effects of higher values of C1S on increased CHD risk, and association with increased plaque vulnerability, along with higher expression in smooth muscle cells and endothelial cells in plaque, supports drug repurposing for management of CHD and atherosclerosis.

We additionally identified four druggable proteins (A2MG, NAGK, PLTP, and SIG14) with associations with CHD and plaque vulnerability. NAGK is involved in amino sugar metabolism and plays a role in atherosclerosis through endothelial cell activation and inflammatory responses. Glucosamine, a substrate of NAGK, has been shown to suppress endothelial cell activation via O-GlcNAc modification, thereby potentially reducing atherosclerotic inflammation(27). By identifying an association of higher values of NAGK with a higher CHD risk and plaque vulnerability, the current study provides further support that inhibiting NAGK may be a viable strategy to treat CHD and atherosclerosis.

Four proteins that are not yet druggable (ARK72, APOF, ATF6B, SWP70) were identified as associated with CHD and plaque vulnerability. Higher values of APOF (apolipoprotein F) were associated with an increased CHD risk and its expression was higher in plaques with higher vulnerability. Previous studies have described APOF as a potential target for novel therapeutic approaches(33). APOF inhibits CETP activity and reduces the transfer of cholesteryl esters from high density lipoprotein to low density lipoprotein (LDL), which is known to reduce LDL cholesterol plasma concentrations and atherosclerosis risk(34, 35). We have previously shown that despite many failed attempts to develop sufficiently potent and selective compounds targeting CETP, the drug target itself remains a viable target for preventing CHD supporting the ongoing phase 3 study of obicetrapib(10, 36, 37).

Despite the robustness of our analyses, our study has a number of potential limitations. Firstly, the GWAS data we sourced used various high throughput assays, varying in accuracy and analytic scope(51). These assays do not directly measure concentrations of proteins or metabolism breakdown products, but evaluate relative values. Therefore, the magnitude of associations is unlikely to be a strong indicator of the effects of pharmacological perturbation of the target, and smaller MR effects may translate towards large drug effects and vice versa. Instead, these results predominantly provide information on the drug mechanisms and whether targets should be inhibited or activated. In the current study, urinary metabolism breakdown products were used as a proxy to identify more distal metabolic processes occurring in, or impacting, atherosclerotic relevant tissues. Hence, the associations between urinary metabolism breakdown products and CHD do not reflect a direct causal mechanism but rather point to the upstream metabolism origins of these breakdown products, which may play an important role in the development of atherosclerosis and the risk of CHD. For some of the MR-prioritised proteins linked with plaque vulnerability, we found an opposite direction of the association of the protein expression in plaque with the plaque vulnerability as compared to the effect of the circulating protein on CHD risk. These findings underscore the complexity of tissue-specific expression, and the molecular pathways involved in atherogenesis. Protein expression patterns can vary by tissue type and stage of atherosclerosis(52, 53), with certain proteins possibly being present due to cell death rather than active secretion, which may affect the interpretation of their direction of effect. Furthermore, MR-prioritised plasma proteins may be involved in CHD through mechanisms in other tissues, such as arterial constriction, without accumulating in plaques. As a result, these proteins may not be represented in our AE findings. However, since plaques do no synthetize proteins but contain proteins originating from plasma, the plaque proteins we identified are likely linked to the plasma proteins prioritised by MR as being related to CHD. Both the GWAS and the AE Biobank data we used predominantly included European individuals, which may limit the generalizability of our findings to non-European populations. In addition, the observational nature of the AE and its high-risk study population introduces a risk of bias due to for example confounding, reverse causation, and index event bias. However, combining these analyses with MR, which is largely confounding-resistant, meaningfully enhances the robustness of our findings.

In conclusion, we identified urinary biomarkers of altered metabolism, predominantly from amino acid metabolism and unclassified origins, associating with CHD risk. By integrating these findings with plasma proteins and plaque vulnerability, we identified 16 plasma proteins as potential drug targets for CHD, supporting the development of novel and repurposed therapeutic strategies. Additionally, our study highlights the value of integrating multi-modal evidence to uncover information potentially relevant for disease diagnosis, prognosis, and etiology.

## Acknowledgements

This research has been conducted using the UK Biobank Resource under Application Number 12113. The authors are grateful to UK Biobank participants.

## Author contributions

SD, EDB, AFS, and SP designed the study. SD and MV accessed the data, verified the data, and performed analyses. SD drafted the manuscript. All authors provided critical input on the analysis, as well as the drafted manuscript.

## Code availability

Analyses were conducted using Python v3.7.13 (for GNU Linux), Pandas v1.3.5, Numpy v1.21.6, bio-misc v0.1.4, and matplotlib v3.4.3. Single-cell RNA sequencing data were analysed in R using Seurat v3(59).

## Data availability

The GWAS data on CHD used in this study can be accessed via https://www.ebi.ac.uk/gwas/studies/GCST90132314 (n=181,522 CHD cases, n=984,168 controls). The GWAS data on urinary metabolism breakdown product values can be accessed via https://www.ebi.ac.uk/gwas/publications/31959995 (n=1,627). The individual GWAS data on plasma protein values can be accessed as follows: deCODE (n=35,559, https://www.decode.com/summarydata/), SCALLOP (n=30,931, https://www.ebi.ac.uk/gwas/publications/33067605), Ahola-Olli *et al*. (n=8,293, https://www.ebi.ac.uk/gwas/publications/27989323), Framingham (n=6,861, https://www.ebi.ac.uk/gwas/publications/21909115), AGES-Reykjavik (n=5,368, https://www.ebi.ac.uk/gwas/publications/35078996), INTERVAL (n=3,301, https://www.ebi.ac.uk/gwas/publications/29875488), Gilly *et al*. (n=1,328, https://www.ebi.ac.uk/gwas/publications/33303764), and Yang *et al*. (n=636, https://www.ebi.ac.uk/gwas/publications/34239129).

