## Supplementary Material for "Integrating multi-modal omics to identify therapeutic atherosclerosis pathways for coronary heart disease"

**Appendix**

**Content**

Appendix Figures S1………………………………………………………………………. 2

Appendix Figures S2………………………………………………………………………. 3

Appendix Figures S3………………………………………………………………………. 4

Appendix Figures S4………………………………………………………………………. 5

Appendix Figures S5………………………………………………………………………. 6

Appendix Figures S6………………………………………………………………………. 7

Supplemental Methods…………………………………………………………………….. 8

Supplemental Results…………………………………………………………………….. 12

References……………………………………………………………………...……… 13

### Appendix Figure S1

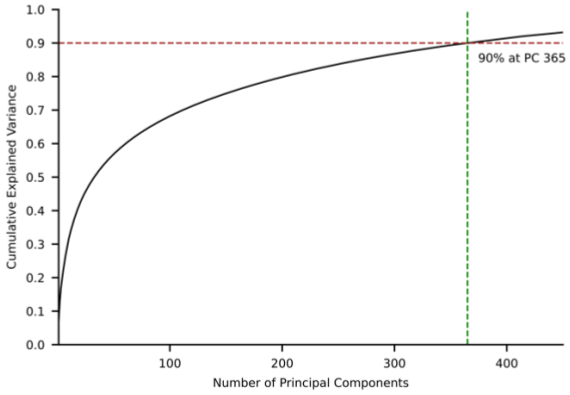


**Figure S1. Cumulative variance of principal component analysis**

N.B. The number of principal components required to explain 90% of the variance of urinary metabolism breakdown product values is depicted. PCA was performed on the Spearman correlation matrix provided by Schlosser *et al*.(12). This study identified and quantified 954 urinary metabolism breakdown products among 1,627 participants.

### Appendix Figure S2

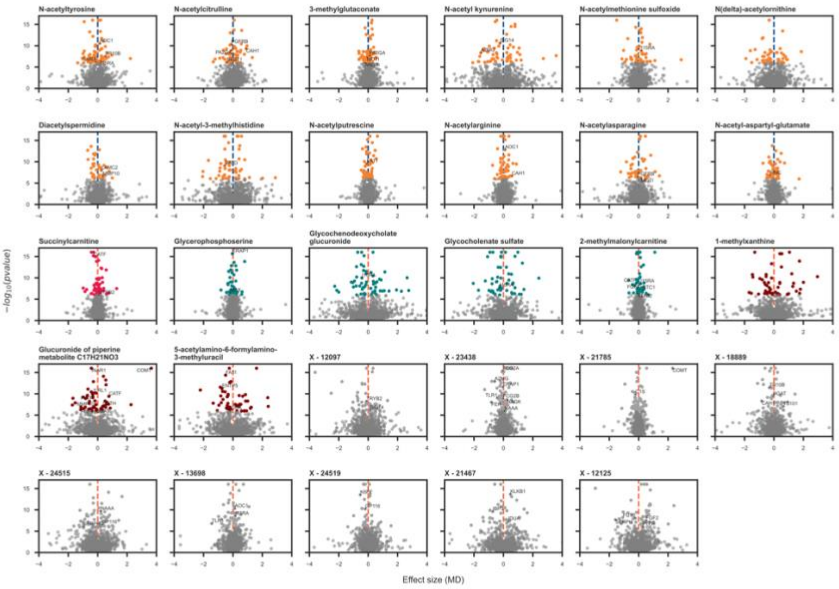


**Figure S2. Volcano plots displaying proteins associated with the metabolism breakdown products identified to be associated with CHD, presented per metabolism breakdown product**

N.B. Labelled proteins are drugged or druggable, which is defined as proteins targeted by an approved compound or by a developmental compound (see Methods). Effect estimates and p-values are obtained from Mendelian randomisation. The p-values have been truncated to -log10 of 16 for visualisation purposes only. Abbreviation: MD = mean difference.

### Appendix Figure S3

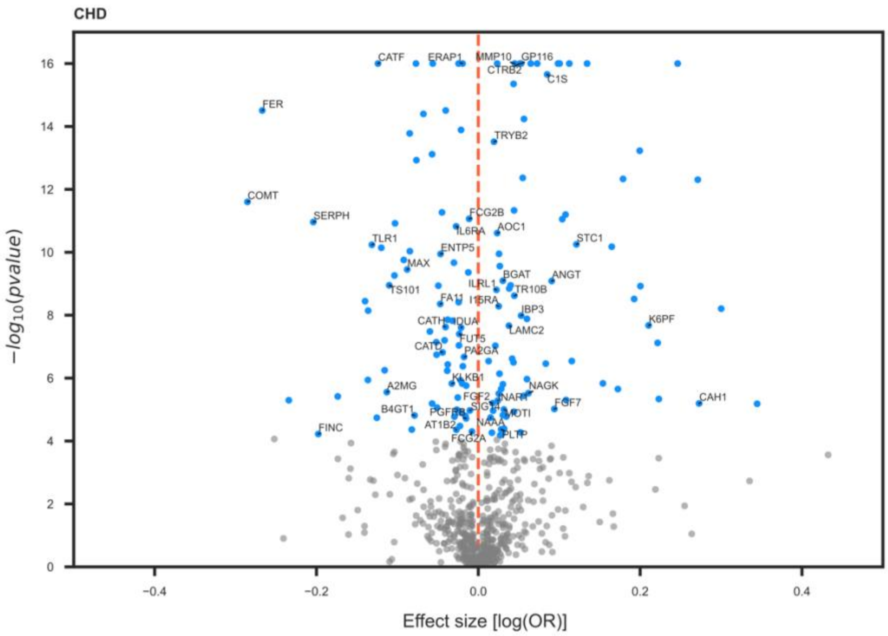


**Figure S3. Volcano plots displaying proteins associated with CHD**

N.B. Labelled proteins are drugged or druggable, which is defined as proteins targeted by a compound or by a developmental compound (see Methods). Effect estimates and p-values are obtained from Mendelian randomisation. The p-values have been truncated to -log10 of 16 for visualisation purposes only. Abbreviations: CHD = coronary heart disease, OR = odds ratio.

### Appendix Figure S4

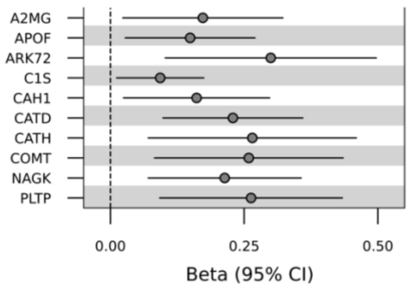


**Figure S4. Associations between protein expression levels and plaque vulnerability index**

N.B. Beta coefficients reflect the increase in normalized count per unit increase in plaque vulnerability index. Protein expression levels and plaque vulnerability index (PVI) data were obtained from carotid plaque samples from patients participating in the Athero-Express Biobank (n=194). Protein expression was measured using Liquid Chromatography-Mass Spectrometry, and PVI is an endpoint ranging from 0 to 5 with one point for plaque characteristics that are considered hallmarks of a vulnerable plaque (moderate/heavy macrophages, no/minor collagen, no/minor smooth muscle cells, lipid core>10% and presence of intraplaque haemorrhage). For more detailed information, please refer to the Methods section and **Appendix Table S7**. Abbreviation: CI = confidence interval.

### Appendix Figure S5

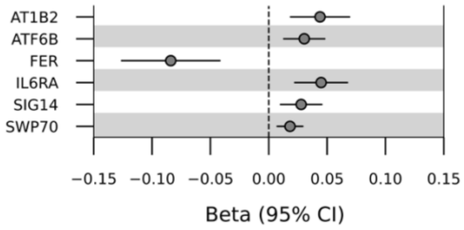


**Figure S5. Associations between mRNA expression levels and plaque vulnerability index**

N.B. Beta coefficients reflect the increase in normalized count per unit increase in plaque vulnerability index. RNA expression levels and plaque vulnerability index (PVI) data were obtained from carotid plaque samples from patients participating in the Athero-Express Biobank (n=632), where mRNA expression was measured using the Illumina NextSeq 500 platform and PVI is an endpoint ranging from 0 to 5 with 1 point for plaque characteristics that are considered hallmarks of a vulnerable plaque (moderate/heavy macrophages, no/minor collagen, no/minor smooth muscle cells, lipid core>10% and presence of intraplaque haemorrhage). For more detailed information, please refer to the Methods section and **Appendix Table S8**. Abbreviation: CI = confidence interval.

### Appendix Figure S6

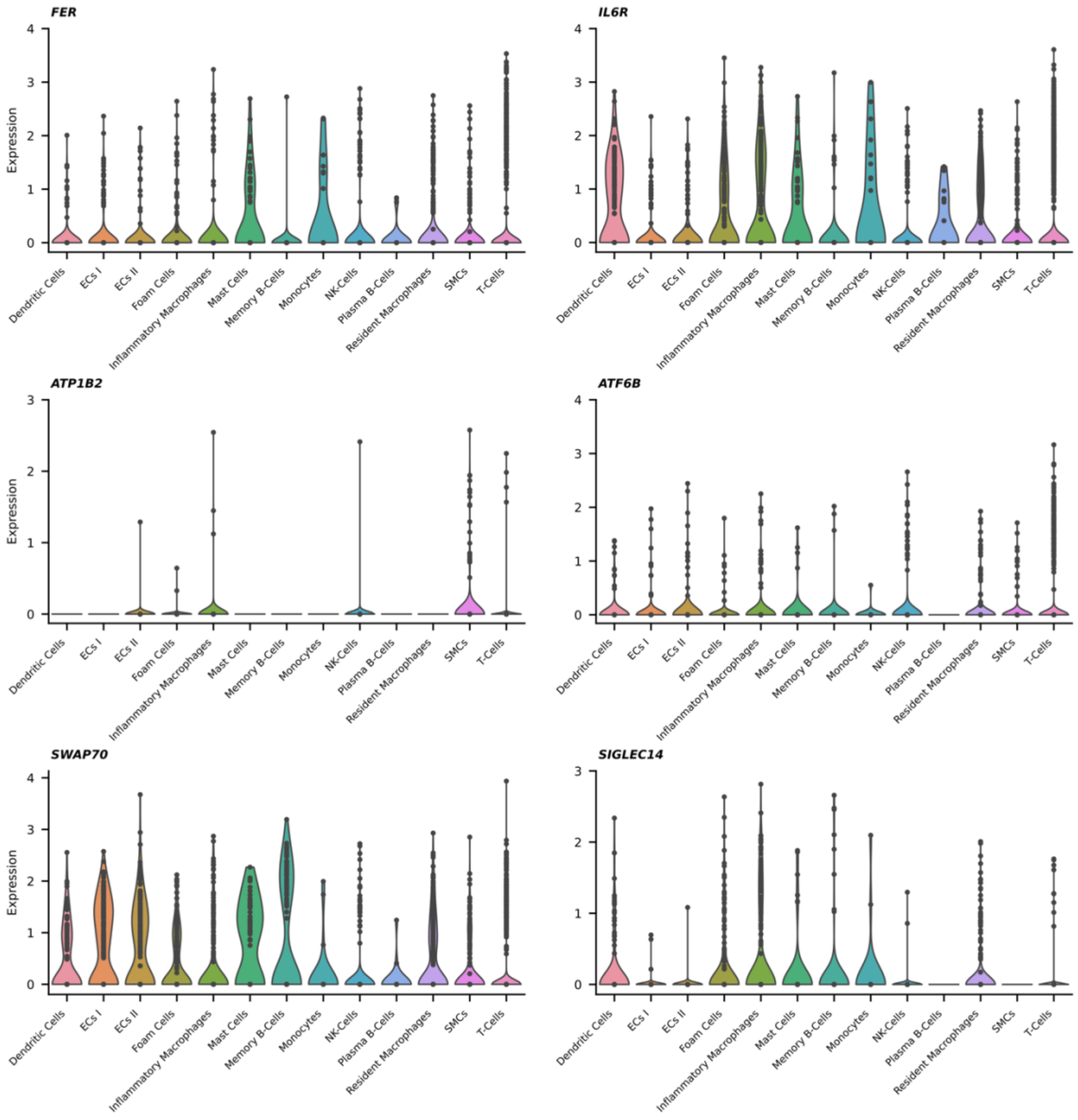


**Figure S6. Distribution of RNA expression of six specific genes across different cell types**

N.B. The presented genes are related to plaque vulnerability in their mRNA expression level in plaque. Single-cell RNA sequencing data were obtained from carotid plaque samples from patients participating in the Athero-Express Biobank (4948 cells, 46 patients). The methods for sample preparation specific to single-cell RNA sequencing are detailed elsewhere(60-62). For more detailed information, please refer to the Methods section. Abbreviations: ECs = endothelial cells, NK-cells = natural killer cells, SMCs = smooth muscle cells.

### Supplemental Methods

*GWAS on CHD*

Genetic associations with CHD were available from Aragam *et al.* (181,522 CHD cases)(13). The GWAS used 10 different studies, where CHD cases were defined using ICD9 and ICD10 codes (I20-I25), covering both coronary artery disease (CAD) as well as more specific conditions like acute coronary syndrome and left main CAD. Definitions varied from including prevalent and incident CAD, additional procedural codes, self-reported CABG, to broader criteria encompassing myocardial infarction, interventions, and ischemic heart disease deaths. The percentage of men across cohorts ranged between 54 and 76%. The average age of cases across cohorts ranged from 34 to 68 years.

*Proteomic GWAS*

Genetic associations with plasma protein values were available from the following eight GWAS: deCODE (SomaLogic assay, n=35,559)(63), SCALLOP (Olink assay, n=30,931)(64), Ahola-Olli et al. (BioRad assay, n=8,293)(65), Framingham (Luminex assay, n=6,861)(66), AGES-Reykjavik (SomaLogic assay, n=5,368)(67), INTERVAL (SomaLogic assay, n=3,301)(68), Gilly et al. (Olink assay, n=1,328)(69) and Yang et al., (SomaLogic assay, n=636)(70).

*Mendelian randomisation assumptions and model selection framework*

MR analysis relies on three core assumptions. First, genetic variants are assumed to be strongly associated with the exposure variable (relevance). Second, it is assumed that genetic variants do not have a common cause with exposure or outcome variables (independence). Third, the effect of genetic variants on the outcome is assumed to be through the exposure variable only (exclusion restriction).

A model selection framework was used to identify the MR model most supported by the data. In the absence of meaningful heterogeneity, the model selection framework favours the IVW method over MR-Egger estimator because it generally yields more accurate estimates (i.e. higher precision) but selects MR-Egger regression in case it provides a demonstratively better fit to the data(17). Within the framework, the goodness of fit to the data is expressed as a Q-statistic.

*Athero-Express Biobank*

We sourced the Athero-Express (AE) Biobank for protein expression levels, mRNA expression levels, single-cell RNA sequencing data, and plaque vulnerability, all measured in carotid plaque samples. The Athero-Express Biobank is an ongoing prospective study of patients undergoing endarterectomy for manifestations of atherosclerosis(71). We had access to clinical data of 2559 patients that underwent carotid endarterectomy. Characteristics of AE patients for who protein expression levels were measured can be found in **Appendix Table S2**. Plaque samples were processed directly after surgery, as described before(71-74). Histological assessment was done according to a standardized protocol. Plaque was divided into segments of 5-mm thickness along the longitudinal axis and the segment with the greatest plaque burden was subjected to histological assessment(71). Semiquantitative estimation of the plaque morphology was performed at ×40 magnification for macrophage infiltration (CD68), SMC content (α-actin), amount of collagen (picrosirius red), and calcification (Hematoxylin-Eosin).

*Plaque vulnerability and histopathological assessment*

Plaque vulnerability index (PVI) was determined for each plaque sample to assess the overall vulnerability of a plaque, and ranges from 0 to 5 with one point for plaque characteristics that are considered hallmarks of a vulnerable plaque: moderate/heavy macrophages, no/minor collagen, no/minor smooth muscle cells, lipid core>10%, and presence of intraplaque haemorrhage, as previously described(75, 76) and based on a previous publication in the Athero-Express(71). The histopathological assessment of carotid plaque samples involved a classification of these hallmarks.

Macrophage infiltration was categorised based on CD68 staining, with criteria distinguishing between 1) absent or minor staining with negative or clusters of fewer than 10 cells and 2) moderate or heavy staining with clusters of more than 10 cells or an abundance of positive cells. Collagen staining was assessed using picrosirius red staining and was categorised as either 1) no or minor staining along the luminal border of the plaque as visualised with and without polarized light or 2) moderate or heavy staining along the entire luminal border of the plaque as visualised with and without polarized light. Smooth muscle cell content was assessed via α-actin staining, distinguishing between 1) no or minor staining across the entire circumference with absent staining at parts of the circumference of the arterial wall and 2) moderate or heaving staining with large number of positive cells along the entire circumference of the arterial wall, with locally at least few scattering cells. The size of the lipid core was estimated visually as a percentage of total plaque area with the use of Hematoxylin-Eosin and picrosirius red stains, with a division into three categories of <10%, 10% to 40% and >40% of the total plaque area, based on the correlation of the lipid core size and plaque stability(77). Plaque haemorrhage was defined as a composite of either bleeding at the luminal side of the plaque as result of plaque disruption(78) or intraplaque haemorrhage which is a haemorrhage within the plaque tissue itself, excluding surgical artifacts such as erythrocyte accumulation along specimen borders. The presence or absence of plaque haemorrhage was examined using Hematoxylin-Eosin and fibrin staining(78, 79).

*Protein and mRNA expression*

Protein expression was measured using Liquid Chromatography-Mass Spectrometry (LC-MS) in carotid plaque samples of 194 Athero-Express Biobank patients (49 women and 145 men). In addition, mRNA expression was measured from 632 samples (156 women and 476 men). As described in more detail before(5), RNA was first isolated from plaque samples using a combination of TriPure reagent, chloroform extraction, and isopropanol precipitation, followed by washing with ethanol. The CEL-seq2 method, known for its high mappability to annotated genes, was chosen for cDNA library preparation, utilizing unique molecular identifiers (UMIs) for precise RNA molecule counting, and the libraries were sequenced on the Illumina NextSeq 500 platform.

*Single-cell RNA sequencing*

Single-cell RNA sequencing data were obtained from 4948 cells and 46 patients (20 women and 26 men) in the Athero-Express Biobank. The methods for sample preparation specific to single-cell RNA sequencing are detailed elsewhere(62). Briefly, viable cells from plaques were sorted individually into wells and immediately frozen at –80°C until further processing using the Sorting and Robot-Assisted Transcriptome Sequencing (SORT-seq) protocol(61). The cells were then pooled into a single library, and the aqueous phase was separated from the oil phase, followed by in vitro transcription and library construction using the CEL-Seq2 protocol(60).

*Determining cellular expression using single-cell RNA sequencing*

An additional test for differential expression using the Wilcoxon rank-sum test was performed for broader clusters of cell types, consisting of three clusters: 1) structural cells comprising endothelial cells I, endothelial cells II, foam cells, and smooth muscle cells, 2) innate immune cells comprising monocytes, resident macrophages, inflammatory macrophages, dendritic cells, mast cells, and NK-cells, and 3) and adaptive immune cells comprising memory B-cells, plasma B-cells, and T-cells.

*Druggability*

Druggability of proteins was obtained from ChEMBL and the British National Formulary (BNF) (step 6 in **Figure 1**). The BNF is a collaborative effort of the British Medical Association and the Royal Pharmaceutical Society that sources information from drug inserts, literature, regulatory agencies, and professional organizations. ChEMBL is a source of information related to clinically used drug targets (from US FDA-approved drugs) and potential targets that are currently under investigation, as described by Finan and colleagues(80).

### Supplemental Results

*Replication of protein effects on CHD*

Of the 113 MR-prioritised proteins, 72 were available in more than one GWAS. Findings were replicated for 60 (83%) protein associations with CHD using a nominal 0.05 p-value threshold. Applying a multiplicity corrected p-value threshold resulted in 42 (58%) replicated protein-CHD associations (**Appendix Table S5**).

*Replication of protein effects on metabolism breakdown products*

Of the 113 MR-prioritised proteins, 67 (59%) were available in more than a single study allowing for MR analyses of protein on urine metabolism breakdown product effects. Metabolism breakdown product associations were replicated for 56 (84%) proteins, when applying a nominal p-value of 0.05. Using a more stringent p-value of $7.46\times10^{-4}$ (0.05 divided by the number of proteins that were available in more than one study) allowed for 45 (67%) replications of protein effects on cardiac outcomes (**Appendix Table S6**).
